# Neural Correlates of Self-Directed Violence: A Large-Sample Resting-State fMRI Study from UK Biobank

**DOI:** 10.1101/2025.05.06.25326773

**Authors:** Jia-Yin Lin, I-Jou Chi, Jun-Ding Zhu, Albert C. Yang

**Affiliations:** Institute of Brain Science, National Yang Ming Chiao Tung University, Taipei, Taiwan; Department of Occupational Therapy, College of Medical Science and Technology, Chung Shan Medical University, Taichung, Taiwan; Occupational Therapy Room, Chung Shan Medical University Hospital, Taichung, Taiwan; Department of Psychiatry, Taipei Veterans General Hospital, Taipei, Taiwan; Department of Medical Research, Taipei Veterans General Hospital, Taipei, Taiwan; School of Medicine, National Yang Ming Chiao Tung University, Taipei, Taiwan; Digital Medicine and Smart Healthcare Research Center, National Yang Ming Chiao Tung University, Taipei, Taiwan

**Keywords:** Suicidal Behavior, Non-Suicidal Self-Directed Violence, Resting-State Functional Connectivity, Limbic System, Reward Circuit

## Abstract

**Importance:** Suicide remains a leading cause of death worldwide, underscoring the importance of identifying neurobiological markers associated with suicidal behaviors. While non-suicidal self-directed violence (NSSDV) is an established risk factor for future suicide attempts (SA), the underlying neural distinctions between these behaviors remain insufficiently understood.

**Objective:** To examine functional connectivity (FC) changes that may differentiate SA from NSSDV based on resting-state functional magnetic resonance imaging (rs-fMRI).

**Design, setting, and participants:** In this case-control study, cross-sectional data of participants with a history of self-directed violence (SDV) were stratified into SA (n = 579) and NSSDV (n = 491) groups based on self-reported questionnaires. Both cross-sectional data and rs-fMRI data were collected from the UK Biobank between October 2016 to January 2024.

**Main outcomes and measures:** FC matrices across different brain regions were analyzed. A general linear regression model was employed to identify significant FC correlations with SA. An exploratory analysis examined the correlation between FC and SDV frequency and interval.

**Results:** A total of 1,070 participants with a history of self-directed violence (SDV) were stratified into SA and NSSDV groups. Compared to NSSDV, the SA group exhibited decreased FC in bilateral amygdala and right putamen, alongside increased FC in the right caudate. Higher frequency of SDV was correlated to increased FC between the orbitofrontal cortex (OFC) and occipital regions, whereas individuals reporting SDV within the past 12 months showed decreased FC between the OFC and both the rectus gyrus and temporal lobe.

**Conclusions and relevance:** This large-sample rs-fMRI study highlights distinct FC alterations associated with different dimensions of self-harm. The limbic and reward circuits appear particularly relevant in differentiating SA from NSSDV and in capturing variations tied to SDV frequency and recency. These insights advance our understanding of suicidal behaviors’ neurobiological underpinnings and may inform targeted risk stratification.

## 1. Introduction

According to the Centers for Disease Control and Prevention (CDC) WISQARS Leading Causes of Death Reports, suicide was the eleventh leading cause of death in the United States in 2022 and exhibited an increasing trend since 2020 (Garnett & Curtin, 2024). The incidence of suicide is complex and multifactorial, influenced by biological, psychological, and socio-environmental factors (Klonsky et al., 2018; Turecki et al., 2019). Prevailing theories attempt to conceptualize suicide attempts (SA), such as the three-step theory and interpersonal theory (Klonsky & May, 2015; Klonsky et al., 2018; Van Orden et al., 2010). Self-harm, defined as self-inflicted injury to one’s own body, regardless of intent, is a significant risk factor for subsequent suicide. Despite self-harm behaviors strongly link to suicide risk, the underlying mechanism and biological evidence remain insufficient (Barredo et al., 2021; Klonsky & May, 2015; Olfson et al., 2017).

Recent studies have reported neurobiological alterations in suicide behaviors, such as monoamine dysregulations and stress-related hypothalamic–pituitary–adrenal axis disturbances (Turecki et al., 2019). Blood-oxygen level dependent (BOLD) signals, extracted from resting-state functional magnetic resonance imaging (rs-fMRI), provide a safe and effective means to observe potential neuronal activity (Fox & Raichle, 2007). A 2019 review summarized structural and functional neuroimaging studies from the past two decades, highlighting the critical role of the cortico-striatal limbic system in suicide behaviors (Schmaal et al., 2020). Despite convergent findings in neuroimaging, larger sample studies are required to validate reliability.

While alterations in resting-state functional connectivity (FC) between individuals with self-harm behaviors have been reported compared to diagnostic controls in different groups, few studies have directly investigated FC differences between SA and NSSI (Huang et al., 2020; Lai et al., 2021; Schmaal et al., 2020). Over the past two decades, suicidal behaviors have been associated with lower FC in the default mode network and reward circuit, and greater FC in the limbic network (S.-G. Kang et al., 2017; Kim et al., 2017; Ordaz et al., 2018; Schmaal et al., 2020). For example, FC alterations in the reward circuit and prefrontal cortex have been identified in individuals with self-harm behaviors and psychiatric disorders (Dimick et al., 2023; Guo et al., 2024). Regarding SA and NSSI differences, Li et al. (2024) investigated FC changes in depressed adolescents with SA and NSSI, indicating divergent inter-network connectivity within visual, limbic, and subcortical networks (Li et al., 2024).

In this study, we hypothesized that alterations in FC exist in individuals with lifetime SA, particularly in the reward circuit and limbic system, compared to those with lifetime non-suicidal self-directed violence (NSSDV), with SA and NSSDV defined according to the CDC’s self-directed violence (SDV) nomenclature recommendations (Crosby et al., 2011). Additionally, most resting-state studies uncovering the biological mechanisms of suicidal behaviors have been limited to small sample sizes (Huang et al., 2020). Participants in this study were therefore selected from the UK Biobank, providing a more comprehensive and robust cohort. The aims of this study were (1) to investigate alterations in FC between lifetime SA and lifetime NSSDV, and (2) to explore FC changes in relation to the frequency and interval of SDV. By expanding understanding of rs-fMRI phenotypes, this research may provide critical insights into the neurobiological distinctions underlying suicidal behaviors, facilitating more precise risk identification and informing targeted early intervention strategies.

## 2. Methods

### 2.1 Participants, Measurement of Frequency and Interval of SDV, and Classification of Suicide Attempts

The UK Biobank is a non-profit, population-based biobank project in the United Kingdom, supported by the Scottish government and various charities. It was made available to registered researchers in 2019 and recruited volunteers aged 40-69 (Miller et al., 2016; Tang, 2018). Demographic data and medical records were collected at enrollment, and mental health web-based questionnaires were regularly administered online. Participants who answered “Yes” to question H3 in the online questionnaire-“Have you deliberately harmed yourself, whether or not you meant to end your life?” - were defined as individuals with lifetime SDV. The frequency of SDV was derived from question H3a: “How many times have you harmed yourself?”. The interval of SDV was obtained from question H3b: “Have you harmed yourself in the last 12 months, whether or not you meant to end your life?”. Participants who answered “Yes” to question H5: “Have you harmed yourself with the intention to end your life?” were classified into the SA group, while those who answered “No” were classified into the NSSDV group. The definition of SDV, SA, and NSSDV followed the CDC’s SDV nomenclature recommendations (Crosby et al., 2011). A total of 6,861 individuals were included in this research. After quality control of available imaging data and exclusion of participants without clinical records, 1,070 individuals were selected for image analysis. Figure 1 presents the flow chart of participant selection. Further details regarding participant selection and UK Biobank questionnaires are described in the supplementary methods.

**Figure 1.**
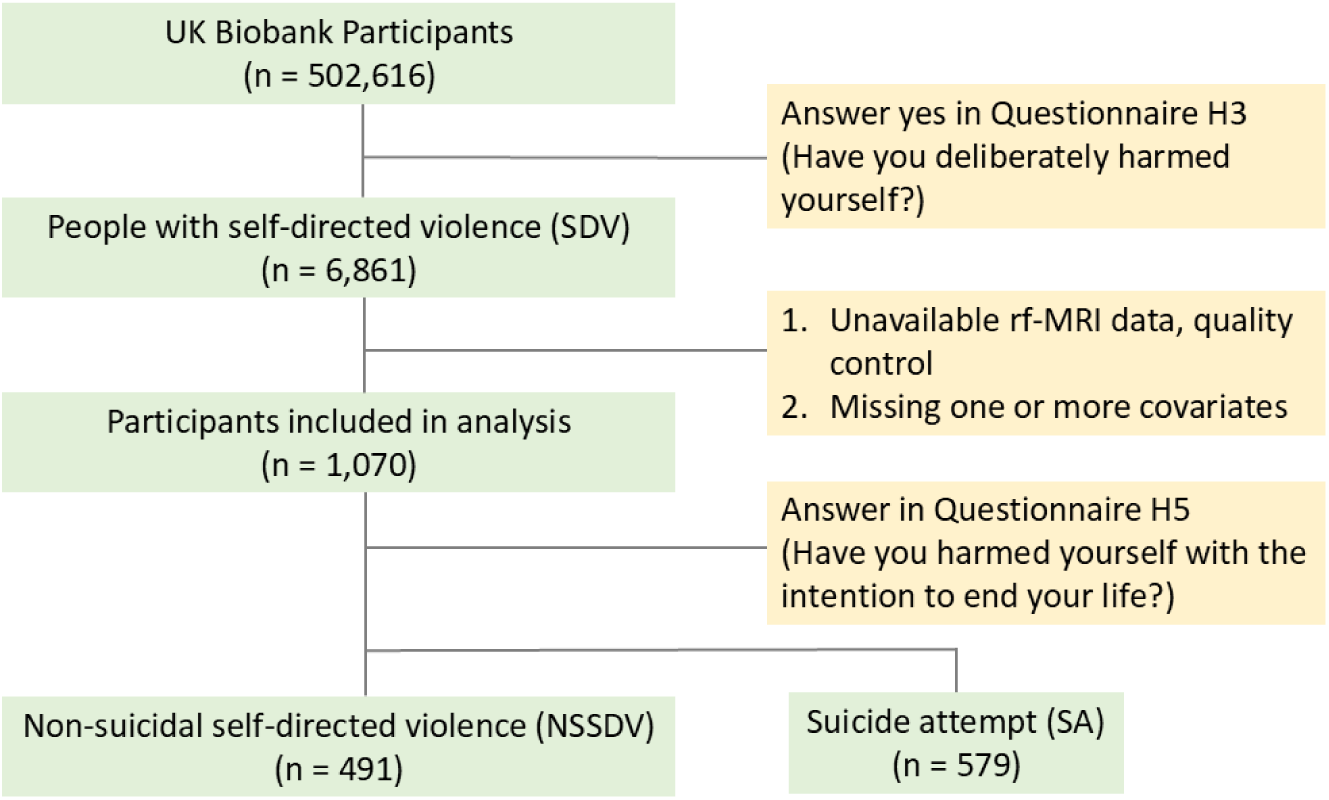
Flowchart of the participant selection. Participants who answered “Yes” in UK biobank online mental health web-based questionnaire H3 was defined as individuals with SDV. Participants who answered “Yes” in UK biobank online mental health web-based questionnaire H5 was defined as SA group, while the one who answered “No” was defined as NSSDV group. Abbreviations: SDV, self-directed violence; SA, suicide attempt; NSSDV, non-suicidal self-directed violence

### 2.2 MRI Acquisition and Preprocessing

Image acquisition and quality control of structural T1-weighted magnetic resonance imaging (T1-weighted MRI) and rs-fMRI were conducted by the UK Biobank team. The images were acquired using the Siemens Skyra 3T running VD13A SP4 with a standard Siemens 32-channel head coil. (Alfaro-Almagro et al., 2018). All images were anonymized using a defacing mask to remove facial structures by the UK Biobank teams. Further scan parameters and details of the image acquisition are presented in Table S1.

Preprocessing was completed using the Data Processing Assistant for rs-fMRI (DPARSF) (Yan et al., 2016) and Statistical Parametric Mapping 12 (SPM12) (Friston, 2007) in the Data Processing & Analysis for Brain Image (DPABI) V4.2_190919 toolbox, an extension in MATLAB (MathWorks, Natick, MA, USA).

Preprocessing of T1-weighted MRI included reorientation along the anterior commissure-posterior commissure line (AC-PC line), regularization to the European template, normalization to MNI 152 space (Montreal Neurological Institute 152 space) (Mazziotta et al., 2001), skull stripping after reorientation, and tissue-type segmentation. Preprocessing of rs-fMRI included the removal of the first 10 time points to correct for initial signal instability, slice timing correction, realignment, reorientation according to the AC-PC line, co-registration to the T1-weighted MRI, normalization to MNI 152 space, head motion correction using the Friston 24-parameter model (Friston et al., 1996), regression of nuisance covariates to remove cardiac and respiratory effects (Hallquist et al., 2013), and band pass filtering performed at 0.01-0.1 Hz.

### 2.3 Functional Connectivity Analysis

After preprocessing, the rs-fMRI data were parcellated into 90 brain regions using the Automated anatomical labelling atlas (AAL-90) (Tzourio-Mazoyer et al., 2002). The time series was extracted from the mean of the blood-oxygen-level-dependent (BOLD) signals in each brain area across the 480 time points, with the interval set as the repetition time (TR = 0.735 sec). Functional connectivity, assessed by calculating Pearson’s correlation between BOLD signals, was defined as the correlation coefficient (r score) for each region pair. Connectivity matrices (resulting in a 90×90 matrix) were constructed, followed by Fisher r-to-z conversion to standardize the r scores to a standard normal distribution (Finn et al., 2015; Rosenberg et al., 2016).

### 2.4 Statistical Analysis

The association between SA, frequency and interval were analyzed using a general linear regression model (GLM). To compare the differences in FC across the 90 brain regions between SA and NSSDV group, the GLM was performed 4,005 times (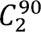 = 4,005), with a statistical significance level set at p < 0.05. The same calculation steps were applied to compare the FC alterations related to the frequency and interval of SDV. Covariates included in the analysis were age, gender, based on prior studies (Alfaro-Almagro et al., 2021; Smith & Nichols, 2018). False discovery rate (FDR) correction was applied for multiple comparison (Benjamini & Hochberg, 1995; Shen et al., 2018).

## 3. Results

### 3.1 Demographic Data

A total of 1,070 participants were included in this study, all of whom had engaged in self-harm at least once. The average age of the participants was 51.82 years (S.D. = 7.08), with 72.71% being female. The average BMI indicated that participants were overweight (mean = 26.59 kg/m^2^, S.D. = 4.81). Additionally, 97.66% of participants identified as non-Hispanic White. Regarding self-harm frequency, approximately half of the participants (n = 532) reported engaging in self-harm only once. In terms of timing, 91.78% (n = 982) indicated that their most recent self-harm incident had occurred more than 12 months ago. 54.11% (n = 579) reported self-harm with the intention of ending their lives, classifying them as the SA group in this study. Demographic characteristics and questionnaire analysis are presented in Table 1.

**Table 1.** Demographic and Self-Harm Characteristics.

| **Characteristics (n=1,070)** | **Value** |
| --- | --- |
| Age, mean (SD), years | 51.82 (7.08) |
| Gender, Female, No. (%) | 778 (72.7%) |
| BMI, mean (SD), kg/m<sup>2</sup> | 26.59 (4.81) |
| Ethnicity |  |
| White | 1045 (97.7%) |
| Mixed | 11 (1.03%) |
| Asian or Asian British | 6 (0.6%) |
| Black or Black British | 4 (0.4%) |
| Chinese | 1 (0.1%) |
| Other | 3 (0.3%) |
| Frequency of SDV, No. (%) |  |
| 1 | 532 (49.7%) |
| 2 | 176 (16.5%) |
| 3 or more | 362 (33.8%) |
| Interval of SDV, No. (%) |  |
| Over 12 months ago | 982 (91.8%) |
| Within the last 12 months | 88 (8.2%) |
| SA, No. (%) |  |
| No | 491 (45.9%) |
| Yes | 579 (54.1%) |

Abbreviations: SDV, self-directed violence; SA, suicide attempt; SD, standard deviation;
BMI, body mass index.

### 3.2 Functional Connectivity Associated with SA

Figure 2 illustrates alterations in FC in the SA group compared to the NSSDV group after FDR correction (q < 0.0498). Individuals with SA exhibited negative correlations in FC between bilateral amygdala and other brain regions, while positive correlations were observed between the right caudate and other brain regions (Figure 2A, Figure S1). Bilateral amygdala, right caudate, and right putamen were the top three brain regions with the most significantly altered FC in SA group (Figure 2B, Table S2). FC between the left insula and right putamen, as well as between the left amygdala and right para-hippocampal gyrus, was negatively correlated with SA when statistical significance was set at p < 0.0005 (Figure 2C). The top fifty significant FC links correlated with SA are presented in Table S5.

**Figure 2.**
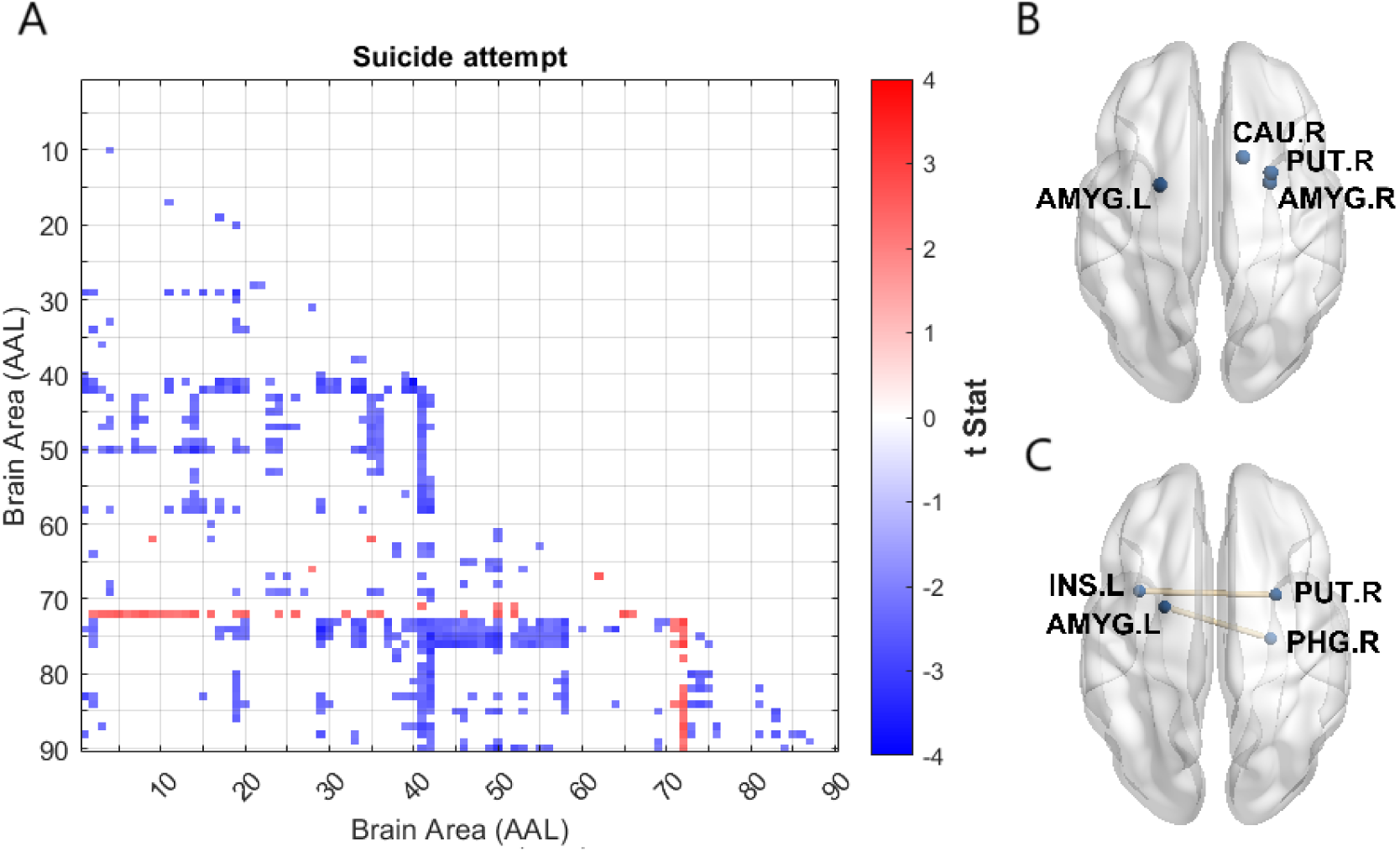
The significant FC correlation in SA group compared to NSSDV group. 486 models were statistically significant (q < 0.0498) after FDR corrections. (A) Negative t (cold color spectrum) value represents SA is associated with lower FC, while positive t value (warm color spectrum) represents the association of higher FC. (B) Top three brain regions with the most statistically significant GLM models, which implied a shift in BOLD dynamics within these regions (C) FC that negatively correlated to SA when setting statistical significance level at p < 0.0005. Abbreviations: FC, functional connectivity; SA, suicide attempt; NSSDV, non-suicidal self-directed violence; FDR, false discovery rate; GLM, general linear regression model; BOLD dynamics, Blood-oxygen level dependent dynamics

### 3.3 Functional Connectivity Associated with Frequency of SDV

Figure 3 illustrates the alterations in FC associated with SDV frequency after FDR correction (q < 0.05). As frequency increased, positive correlations in FC were observed. The results showed clusters of positive correlation between bilateral middle frontal gyrus, orbitofrontal cortex (OFC) and cuneus, occipital lobes (Figure 3A, Figure S2). The left inferior orbitofrontal gyrus, right cuneus and right inferior occipital gyrus were the top three brain regions with the most significantly altered FC following increased SDV frequency (Figure 3B, Table S3). FC between the right cuneus and right OFC was positively correlated with SDV frequency when statistical significance was set at p < 0.0001 (Figure 3C). The top fifty significant FC links correlated with SDV frequency are presented in Table S6.

**Figure 3.**
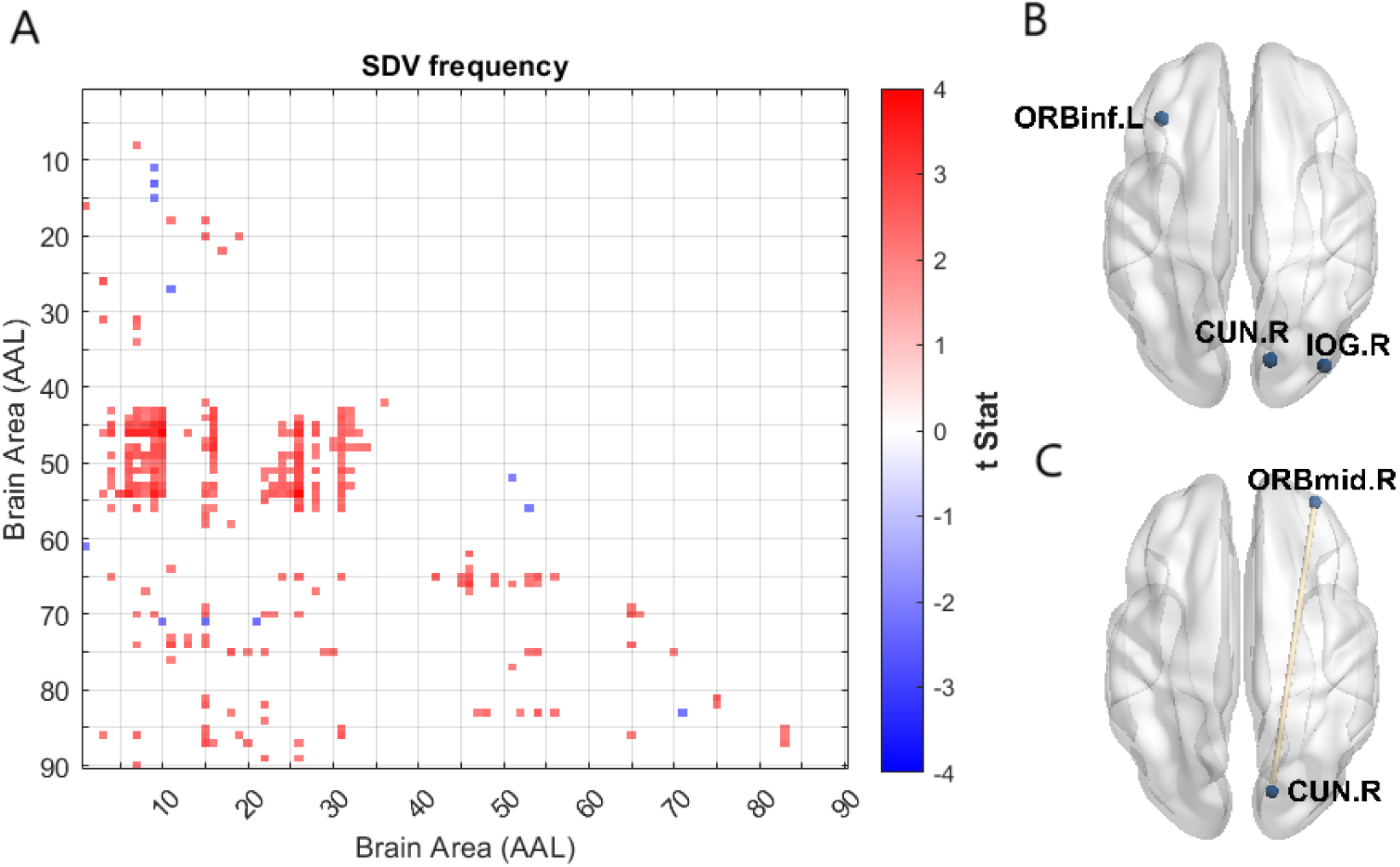
The correlation between frequency of SDV and FC. 256 models were statistically significant (q < 0.0500) after FDR corrections. (A) Positive t value (warm color spectrum) represents frequency of SDV is associated with higher FC, while negative t value (cold color spectrum) represents the association of lower FC. (B) Top three brain regions with the most significant GLM model, which implied the shift in BOLD dynamics within these regions (C) FC that positively correlated to SDV frequency when setting statistical significance level at p < 0.0001. Abbreviations: SDV, self-directed violence; FC, functional connectivity; FDR, false discovery rate; GLM, general linear regression model; BOLD dynamics, Blood-oxygen level dependent dynamics

### 3.4 Functional Connectivity Associated with Self-harmed Interval

Figure 4 depicts alterations in FC associated with the SDV interval after FDR correction (q < 0.0498). The results showed that individuals who engaged in SDV within the past 12 months exhibited negative correlations in FC between bilateral middle frontal gyrus, OFC and rectus gyrus, temporal lobe (Figure 4A, Figure S3). The right middle orbitofrontal gyrus, right superior orbitofrontal gyrus, and right middle frontal gyrus were the top three brain regions with the most significantly altered FC in individuals with SDV within the past 12 months (Figure 4B, Table S4). FC between the right anterior cingulate gyrus and inferior temporal gyrus, as well as between the right lingual gyrus and left calcarine cortex, was negatively correlated with the SDV interval when statistical significance was set at p < 0.001 (Figure 4C). The top fifty significant FC links correlated with the SDV interval are presented in Table S7.

**Figure 4.**
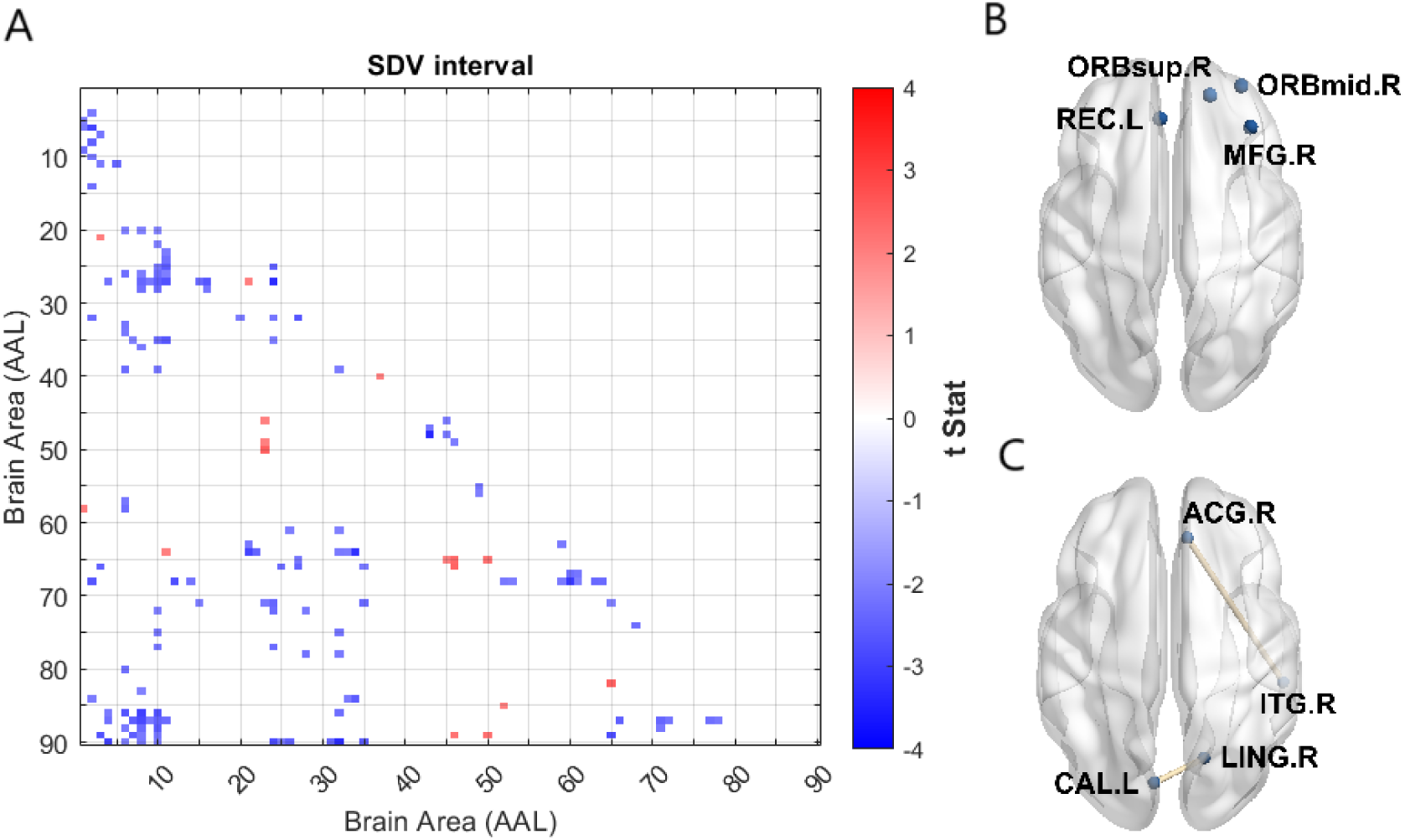
The correlation between intervals of SDV and FC. 155 models were statistically significant (q < 0.0498) after FDR corrections. (A) Negative t value (cold color spectrum) represents SDV interval was associated with lower FC, while positive t value (warm color spectrum) represents the association of higher FC. (B) Top three brain regions with the most significant GLM models, which implied the shift in BOLD dynamics within these regions (C) FC that negatively correlated to SDV interval when setting statistical significance level at p < 0.001. Abbreviations: SDV, self-directed violence; FC, functional connectivity; FDR, false discovery rate; GLM, general linear regression model; BOLD dynamics, Blood-oxygen level dependent dynamics

## 4. Discussion

This study explored FC differences in suicidal behaviors among adults. The key FC difference identified in this large-sample database study reflect altered reward circuits and limbic systems associated with clinical manifestations in individuals with SDV. First, widespread FC alterations in the bilateral amygdala and right caudate were observed in the SA group compared to NSSDV group. Second, a significant increase in FC between the OFC and occipital lobes was observed with higher SDV frequency. Third, decreased FC between the OFC and rectus gyrus, as well as OFC and temporal lobe, was found in individuals who engaged in SDV within the past 12 months.

The connectivity alterations identified in this study support neurofunctional distinctions between SA and NSSI. Although both SA and NSSI are often driven by a desire to relieve distress, they differ in suicidal intent, lethality, and interpersonal factors (Hamza et al., 2012; Klonsky & May, 2015). Previous studies suggest that DMN, limbic network, and reward network play key roles in SA among adults (S.-G. Kang et al., 2017; Kim et al., 2017; Ordaz et al., 2018; Schmaal et al., 2020), while neuroimaging findings specific to NSSI alone have been less reported (Domínguez-Baleón et al., 2018). The observed FC alterations align with prior studies suggesting limbic network dysfunction and reward circuit differences (Huang et al., 2020; S.-G. Kang et al., 2017; Myung et al., 2016).

While sample size is crucial for reliability, analytic methodology also influences connectivity results. Matthew Thompson et al. (2023) examined triple-network differences between individuals with SA and those with self-directed violence using the same dataset, finding no significant differences in connectivity in extracted ICA components (Jenkinson et al., 2002; Thompson et al., 2023). ICA requires manual identification of neuroanatomical relevance (Fox & Raichle, 2007), and the limited number of components may overlook key systems. In contrast, our study accounted for physical artifacts through nuisance regression and used the AAL atlas for region-based connectivity analysis.

A negative FC association in the bilateral amygdala and a positive association in the right caudate was observed in individuals with SA compared to NSSDV. The amygdala is a crucial structure within the limbic network, involved in processing emotions such as fear, anxiety, and aggression (Torrico & Abdijadid, 2025). Dysregulation in amygdala neurotransmission has been strongly linked to anxiety disorders (Babaev et al., 2018), a well-established personality trait associated with suicidal behavior (Turecki et al., 2019). Previous studies have reported decreased left amygdala FC with the left lateral occipital cortex in adolescent with bipolar disorder and self-harm (Dimick et al., 2023), as well as increased amygdala connectivity with the insula, superior OFC and middle temporal cortex in adults with major depressive disorder (MDD) and SA (S. G. Kang et al., 2017).

The caudate contains the nucleus accumbens, a core hub of reward circuit, and plays a key role in reward processing, motivation, and emotion (Driscoll et al., 2025; Haber & Knutson, 2010). Research has highlighted the reward circuit as a key component in impulsive regulation, with dopaminergic dysregulation implicated in reward deficiency syndromes (Basar et al., 2010; Blum et al., 2000). A review found evidence of blunted striatal activation in individuals with suicidal thoughts, behaviors and NSSI (Auerbach et al., 2021). Given the stronger association between suicidal behavior and impulsive-aggressive traits (Turecki et al., 2019), the increased connectivity in the caudate observed in this study may reflect dysregulation of the reward circuit in individuals with SA.

A positive association of FC in the left OFC was observed with increased SDV frequency, while a negative association in right OFC was found in individuals with a shorter interval since their last SDV. The OFC plays a key role in decision-making, emotion regulation, and response inhibition and is functionally connected to both the limbic network and the reward circuit (Haber & Knutson, 2010; Kringelbach, 2005; Rolls, 2015). Self-harm is often viewed as a maladaptive coping strategy when individuals experience stress (Hamza et al., 2012; Turecki et al., 2019). Dysfunctional OFC activity may lead individuals to resort to self-harm as an ineffective stress-relief strategy by impairing positive thinking and response inhibition. Task-based fMRI studies have identified increased FC between the OFC and precuneus, supplementary motor areas during reward processing in individuals with NSSI (Lin et al., 2024). Guo et al. (2024) found a negative correlation between FC in the right OFC and right insula with NSSI frequency in adolescents with depression, consistent with the findings of this study (Guo et al., 2024).

The increased FC associated with SDV frequency and visual-related regions, including cuneus and inferior occipital gyrus, has been less frequently discussed in previous neuroimaging studies on suicidal behavior (Schmaal et al., 2020). Regions in occipital lobes contribute to the perception of emotional information through pathways from visual cortex to OFC (Fusar-Poli et al., 2009; Rolls, 2015). The positive correlation between FC and SDV frequency may suggest the role of visual stimuli in the neurocircuitry underlying SDV. A facial expression recognition study reported that adolescents with NSSI rated emotional stimuli as more unpleasant and arousing compared to controls (In-Albon et al., 2015). In rs-fMRI research, divergent alterations in visual network were observed in depressed adolescents engaging in NSSI and SA (Li et al., 2024).

Several limitations should be considered when interpreting these findings. First, while UK Biobank provides extensive and valuable data, distinguishing subgroups within the psychiatric population at high risk of suicidal behaviors remains challenging. Up to 6,000 participants without a psychiatric diagnosis engaged in self-directed violence, highlighting the underdiagnosis of mental illness in the general population. According to the CDC, more than half of individuals who died by suicide had no formal psychiatric diagnosis at the time of their death (Control & Prevention, 2018). Second, there is inherent heterogeneity in medical history and clinical profile, including factors such as early life adversity and substance use. However, this heterogeneity reflects a real-world population, enhancing the generalizability of the findings. Third, the cross-sectional design of this study prevents causal inference between FC alterations and suicidal behaviors. Longitudinal studies are needed to establish causality, though they face challenges due to the complex and unpredictable nature of suicide risk factors (Klonsky & May, 2015).

## 5. Conclusions

This study leveraged the large-sample UK Biobank to investigate differences in FC between individuals with lifetime SA and those with lifetime NSSDV. The main findings revealed that SA was associated with decreased FC in the bilateral amygdala and right putamen, alongside increased FC in the right caudate. Higher SDV frequency correlated with increased FC between the OFC and visual-related occipital lobes, while recent SDV within the past 12 months was linked to decreased FC in the OFC, particularly with the rectus gyrus and temporal lobe. These observations support the notion of neurobiological dysregulation in suicidal behaviors, especially in limbic system–associated brain regions. Further research is needed to examine how predisposition factors (e.g., early-life adversity, family history) influence functional neuroimaging findings. Overall, this study offers valuable insights into the neurobiological mechanisms underlying suicidal behaviors and may aid in risk stratification, ultimately informing public suicide prevention efforts.

## Data Availability

This research has been conducted using the UK Biobank Resource under Application Number 91995. All data produced in the present study are available to bona fide researchers upon application to UK Biobank.

https://www.ukbiobank.ac.uk/

## Acknowledgements

This study used data from UK Biobank under application number 91995. We thank to UK Biobank Access, the epidemiological and data analyst team, for the tireless work to help researchers access the data. We would particularly like to thank the 500,000 participants in the UK Biobank study for their enormous generosity and altruism and their continued interest, support, and involvement.

