## Supplemental Materials for "Neural Correlates of Self-Directed Violence: A Large-Sample Resting-State fMRI Study from UK Biobank"

**Running Title**: Neural Connectivity in Suicidal vs. Non-Suicidal Self-Harm

**Author Affiliation:**

Jia-Yin Lin^1^, I-Jou Chi^1^, Jun-Ding Zhu^2,3^, Albert C. Yang^1,4,5,6,7*^

^1^ Institute of Brain Science, National Yang Ming Chiao Tung University, Taipei, Taiwan

^2^ Department of Occupational Therapy, Chung Shan Medical University, Taichung, Taiwan

^3^ Occupational Therapy Room, Chung Shan Medical University Hospital, Taichung, Taiwan

^4^ Department of Psychiatry, Taipei Veterans General Hospital, Taipei, Taiwan

^5^ Department of Medical Research, Taipei Veterans General Hospital, Taipei, Taiwan

^6^ School of Medicine, National Yang Ming Chiao Tung University, Taipei, Taiwan

^7^ Digital Medicine and Smart Healthcare Research Center, National Yang Ming Chiao Tung University, Taipei, Taiwan

***Corresponding Author:** Dr. Albert C. Yang, M.D., Ph.D.

Present address: No. 155 Sec. 2 Linong St., Beitou Dist., Taipei City, Taiwan 112, National Yang Ming Chiao Tung University

Telephone number: +886228267000#66555

**Supplemental Methods**

**Participants**

The UK Biobank, which includes 500,000 participants, was made available to registered researchers in 2019 (Tang, 2018). Demographic data and medical records were collected at the time of enrollment, and mental health questionnaires were regularly administered online. Participants who answer “Yes” to question H3 in the online mental health web-based questionnaire: “Have you deliberately harmed yourself, whether or not you meant to end your life?” (UK Biobank field ID 20480) aligned with the SDV definition in CDC’s SDV nomenclature recommendations (Crosby et al., 2011). A total of 6,861 individuals were included in this research. After quality control of available imaging data and the exclusion of participants without clinical records, 1,070 individuals were selected for image analysis.

**Measurement of Frequency and Interval of Self-Directed Violence (SDV), and Classification of Suicide Attempts**

The frequency and interval of SDV for each participant and the SA group were obtained from the section on harm behaviors in the mental health web-based questionnaires of the UK Biobank. This questionnaire was designed by the UK Biobank working groups to access details of self-harm behaviors. The frequency of SDV was derived from question H3a: “How many times have you harmed yourself?” (UK Biobank field ID 20482), with selectable responses: “1”, “2”, “3 or more”, and “Prefer not to answer”. The interval of SDV was obtained from question H3b: “Have you harmed yourself in the last 12 months, whether or not you meant to end your life?” (UK Biobank field ID 20481), with selectable responses: “No”, “Yes”, and “Prefer not to answer”. Participants who answered “Yes” to question H5: “Have you harmed yourself with the intention to end your life?” (UK Biobank field ID 20483) were classified into the SA group, while those who answered “No” were classified into the NSSDV group. The definition of SA and NSSDV aligned with the CDC’s SDV nomenclature recommendations (Crosby et al., 2011). The selectable responses for question H5 included “No”, “Yes”, and “Prefer not to answer”. Further details on the questionnaires can be accessed through the UK Biobank mental health web-based questionnaires protocol (<https://biobank.ctsu.ox.ac.uk/crystal/refer.cgi?id=22>) on the internet.

**Supplemental Figures and Tables**

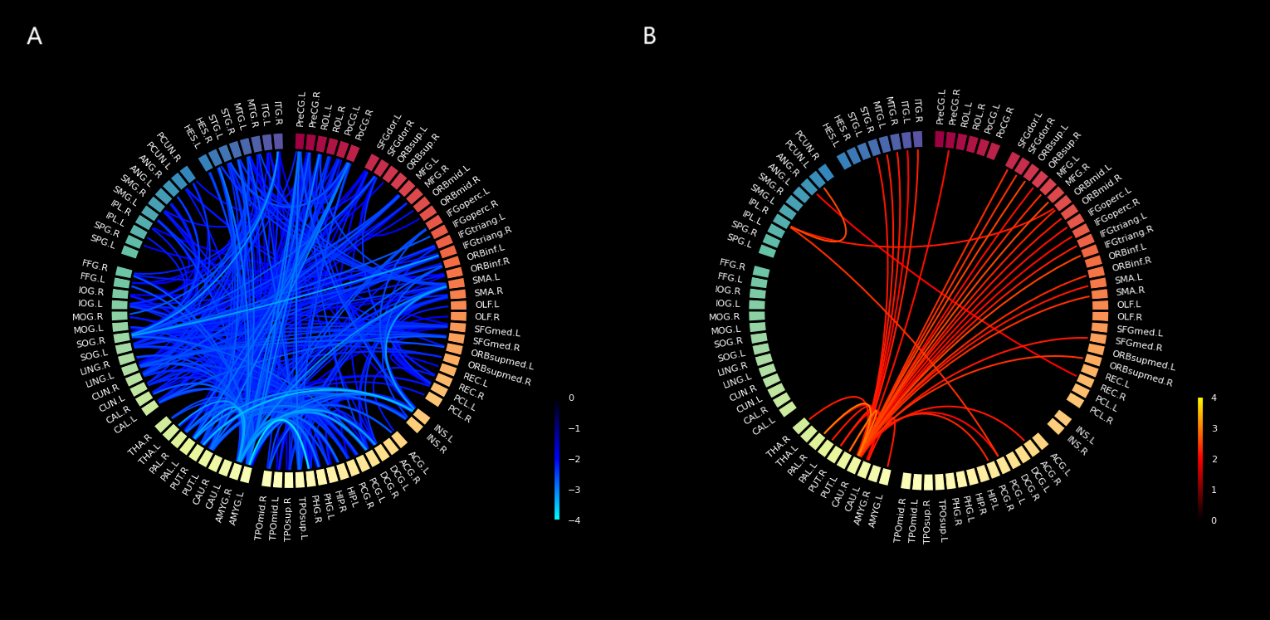
**Figure S1.** Functional connectivity with significant GLM models after FDR correction (q < 0.0498) in individual with SA compared to NSSDV. The spectrum indices represent the t-values of FC correlations in the GLM models. Positive t-values are depicted in a warm spectrum, while negative t-values are shown in a cold spectrum. (A) Widespread negative FC correlations were observed in bilateral amygdala. (B) A positive FC correlation was observed in the right caudate. Models were adjusted for gender and age. Abbreviations: GLM, general linear regression model; FDR, false discovery rate; SA, suicide attempt; NSSDV, non-suicidal self-directed violence; FC, functional connectivity

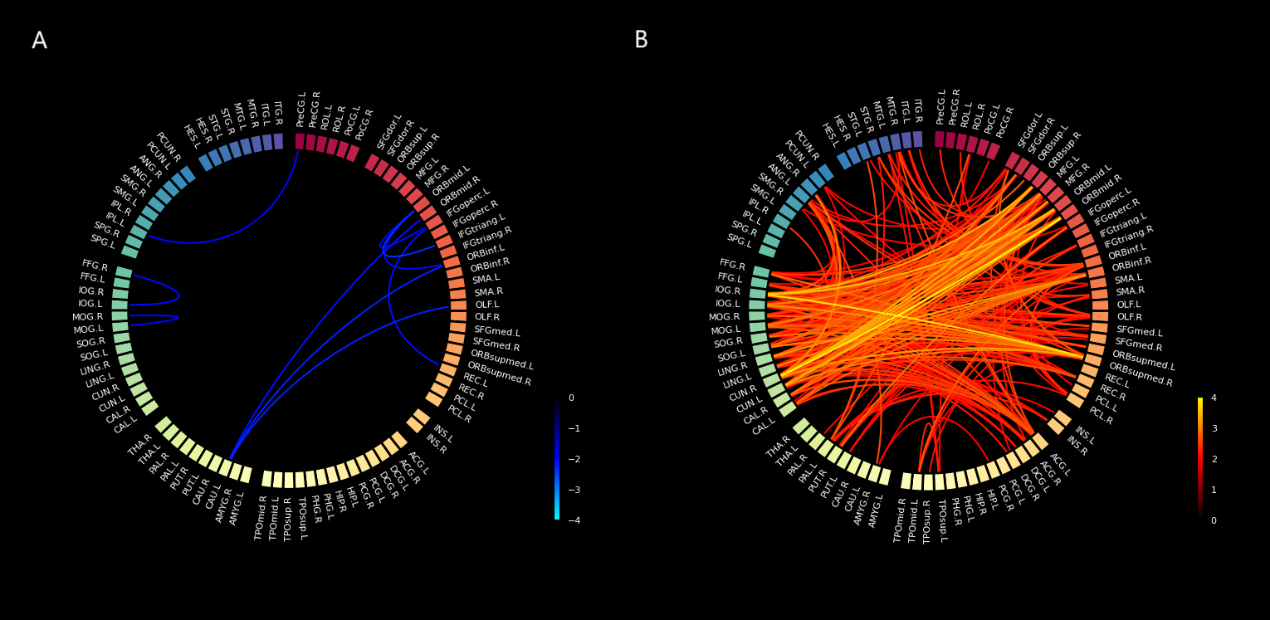

**Figure S2.** Functional connectivity with significant GLM models after FDR correction (q < 0.0500) associated with SDV frequency. The spectrum indices represent the t-values of FC correlations in the GLM models. Positive t-values are depicted in a warm spectrum, while negative t-values are shown in a cold spectrum. The results showed positive FC correlations between the orbitofrontal cortex (OFC) and occipital lobes, as illustrated in Figure S2B. Models were adjusted for gender and age. Abbreviations: GLM, general linear regression model; FDR, false discovery rate; SDV, self-directed violence; FC, functional connectivity

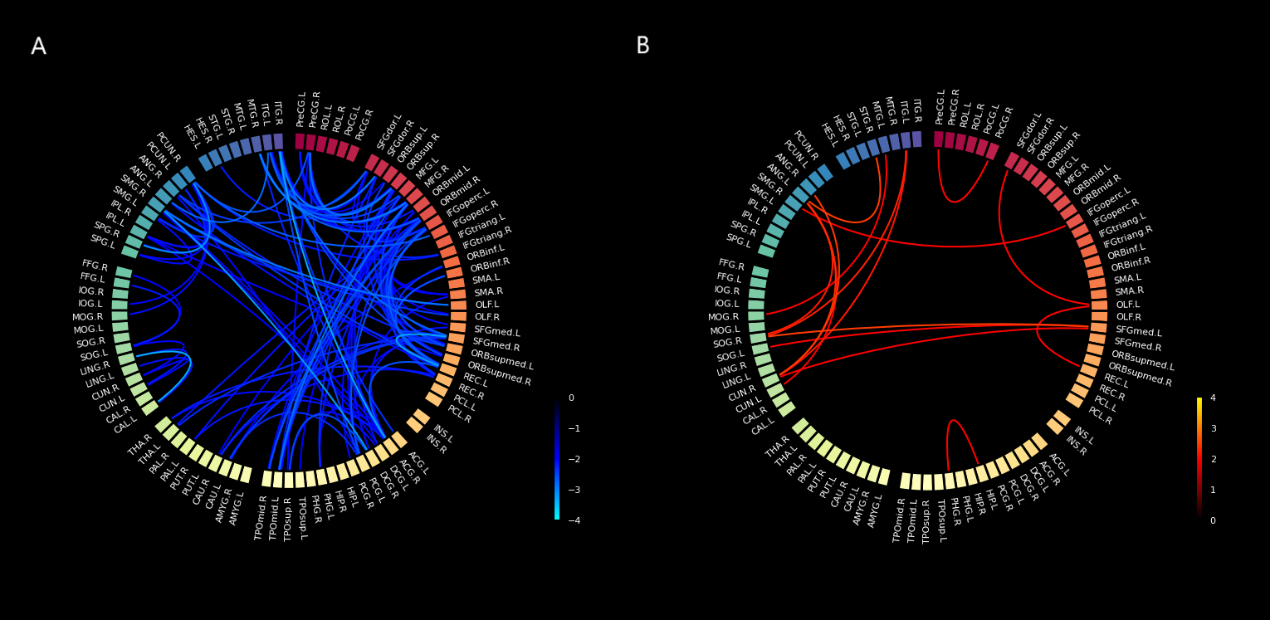

**Figure S3.** Functional connectivity with significant GLM models after FDR correction (q < 0.0498) associated with SDV interval. The spectrum indices represent the t-values of FC correlations in the GLM models, where positive t-values are depicted in a warm spectrum and negative t-values in a cold spectrum. Negative FC correlations were observed between the OFC and rectus gyrus, as well as between the OFC and temporal lobe, as shown in Figure S3A. Models were adjusted for gender and age. Abbreviations: GLM, general linear regression model; FDR, false discovery rate; SDV, self-directed violence; FC, functional connectivity; OFC, orbitofrontal cortex

**Table S1.** Detailed MRI scan parameters. T1-weighted MRI was acquired using 3-D magnetization prepared rapid gradient echo imaging (3D MPRAGE). Rs-fMRI was acquired using gradient echo-planar imaging with ×8 multislice acceleration (GE-EPI with ×8 multislice acceleration) (Alfaro-Almagro et al., 2018). Details of the image acquisition are provided in the UK Biobank data acquisition and preprocessing protocol (<https://www.fmrib.ox.ac.uk/ukbiobank/protocol/V4_23092014.pdf>) and its associated documents (<https://biobank.ctsu.ox.ac.uk/crystal/crystal/docs/brain_mri.pdf>).

| **Imaging Type** | **Parameters** |
| --- | --- |
| T1-weighted MRI | TI (inverse time) = 880 ms, TE (echo time) = 2.01 ms,  FOV (field of view) = 208×256×256 matrix,  Resolution = 1.0×1.0×1.0 mm, FA (flip angle) = 8° |
| Resting-state fMRI | TR (repetition time) = 735 ms, TE (echo time) = 39 ms,  FOV (field of view) = 88×88×64 matrix,  Resolution = 2.4×2.4×2.4 mm, FA (flip angle) = 52°,  Scan duration = 6 min, Time point = 490 |

**Table S2.** Top three brain regions most frequently involved in significant GLM models in SA group. Abbreviations: GLM, general linear regression model; SA, suicide attempt; AAL, Automated anatomical labelling atlas

| **Brain regions** | **AAL** | **Frequency in significant GLM models** |
| --- | --- | --- |
| L. Amygdala | 41 | 54 |
| R. Amygdala | 42 | 48 |
| R. Caudate nucleus | 72 | 40 |
| R. Lenticular nucleus, putamen | 74 | 40 |

**Table S3.** Top three brain regions most frequently involved in significant GLM models correlated to SDV frequency. Abbreviations: GLM, general linear regression model; SDV, self-directed violence; AAL, Automated anatomical labelling atlas

| **Brain regions** | **AAL** | **Frequency in significant GLM models** |
| --- | --- | --- |
| L. Inferior frontal gyrus, orbital part | 15 | 22 |
| R. Cuneus | 46 | 21 |
| R. Inferior occipital gyrus | 54 | 20 |

**Table S4.** Top three brain regions most frequently involved in significant GLM models correlated to SDV interval. Abbreviations: GLM, general linear regression model; SDV, self-directed violence; AAL, Automated anatomical labelling atlas

| **Brain regions** | **AAL** | **Frequency in significant GLM models** |
| --- | --- | --- |
| R. Middle frontal gyrus, orbital part | 10 | 17 |
| R. Superior frontal gyrus, orbital part | 6 | 13 |
| R. Middle frontal gyrus | 8 | 12 |
| L. Gyrus rectus | 27 | 12 |

**Table S5.** Top 50 significant FC correlations in SA groups are presented below, summarized from Figure 2. A total of 486 models remained statistically significant (q < 0.0498) after FDR correction. A positive t-value indicates increased FC in the SA group, while a negative t-value indicates decreased FC. Abbreviations: FC, functional connectivity; SA, suicide attempt; FDR, false discovery rate

| **Label_1** | **Region_1** | **Label_2** | **Region_2** | **p_value** | **t_value** |
| --- | --- | --- | --- | --- | --- |
| 41 | L. Amygdala | 40 | R. Parahippocampal gyrus | 0.000149 | -3.80683 |
| 74 | R. Lenticular nucleus, putamen | 29 | L. Insula | 0.000488 | -3.49845 |
| 75 | L. Lenticular nucleus, pallidum | 42 | R. Amygdala | 0.000737 | -3.38552 |
| 42 | R. Amygdala | 33 | L. Median cingulate and paracingulate gyri | 0.001119 | -3.26771 |
| 50 | R. Superior occipital gyrus | 14 | R. Inferior frontal gyrus, triangular part | 0.001133 | -3.26429 |
| 29 | L. Insula | 19 | L. Supplementary motor area | 0.001211 | -3.24519 |
| 42 | R. Amygdala | 34 | R. Median cingulate and paracingulate gyri | 0.001239 | -3.23856 |
| 77 | L. Thalamus | 42 | R. Amygdala | 0.00146 | -3.19092 |
| 75 | L. Lenticular nucleus, pallidum | 50 | R. Superior occipital gyrus | 0.001712 | -3.14417 |
| 90 | R. Inferior temporal gyrus | 50 | R. Superior occipital gyrus | 0.001758 | -3.13646 |
| 42 | R. Amygdala | 40 | R. Parahippocampal gyrus | 0.001892 | -3.11459 |
| 75 | L. Lenticular nucleus, pallidum | 29 | L. Insula | 0.001975 | -3.10171 |
| 42 | R. Amygdala | 19 | L. Supplementary motor area | 0.002229 | -3.06549 |
| 76 | R. Lenticular nucleus, pallidum | 50 | R. Superior occipital gyrus | 0.002452 | -3.03655 |
| 58 | R. Postcentral gyrus | 42 | R. Amygdala | 0.002455 | -3.03624 |
| 76 | R. Lenticular nucleus, pallidum | 45 | L. Cuneus | 0.002455 | -3.03623 |
| 74 | R. Lenticular nucleus, putamen | 64 | R. Supramarginal gyrus | 0.002629 | -3.01534 |
| 76 | R. Lenticular nucleus, pallidum | 72 | R. Caudate nucleus | 0.002973 | 2.977524 |
| 76 | R. Lenticular nucleus, pallidum | 43 | L. Calcarine fissure and surrounding cortex | 0.00301 | -2.97364 |
| 76 | R. Lenticular nucleus, pallidum | 44 | R. Calcarine fissure and surrounding cortex | 0.003099 | -2.96466 |
| 73 | L. Lenticular nucleus, putamen | 58 | R. Postcentral gyrus | 0.003155 | -2.95912 |
| 41 | L. Amygdala | 17 | L. Rolandic operculum | 0.003187 | -2.9559 |
| 74 | R. Lenticular nucleus, putamen | 34 | R. Median cingulate and paracingulate gyri | 0.003233 | -2.95144 |
| 41 | L. Amygdala | 29 | L. Insula | 0.003406 | -2.93519 |
| 75 | L. Lenticular nucleus, pallidum | 49 | L. Superior occipital gyrus | 0.003435 | -2.93253 |
| 42 | R. Amygdala | 39 | L. Parahippocampal gyrus | 0.003515 | -2.92533 |
| 76 | R. Lenticular nucleus, pallidum | 49 | L. Superior occipital gyrus | 0.003565 | -2.92092 |
| 41 | L. Amygdala | 1 | L. Precentral gyrus | 0.003717 | -2.90777 |
| 73 | L. Lenticular nucleus, putamen | 72 | R. Caudate nucleus | 0.003834 | 2.898008 |
| 80 | R. Heschl gyrus | 42 | R. Amygdala | 0.003886 | -2.89371 |
| 87 | L. Temporal pole: middle temporal gyrus | 41 | L. Amygdala | 0.003925 | -2.89058 |
| 58 | R. Postcentral gyrus | 41 | L. Amygdala | 0.003929 | -2.89023 |
| 75 | L. Lenticular nucleus, pallidum | 58 | R. Postcentral gyrus | 0.003983 | -2.88587 |
| 76 | R. Lenticular nucleus, pallidum | 46 | R. Cuneus | 0.004014 | -2.8834 |
| 41 | L. Amygdala | 34 | R. Median cingulate and paracingulate gyri | 0.004124 | -2.87487 |
| 42 | R. Amygdala | 1 | L. Precentral gyrus | 0.004238 | -2.86613 |
| 76 | R. Lenticular nucleus, pallidum | 42 | R. Amygdala | 0.004358 | -2.85726 |
| 83 | L. Temporal pole: superior temporal gyrus | 1 | L. Precentral gyrus | 0.004487 | -2.8479 |
| 76 | R. Lenticular nucleus, pallidum | 47 | L. Lingual gyrus | 0.004863 | -2.82196 |
| 73 | L. Lenticular nucleus, putamen | 37 | L. Hippocampus | 0.00497 | -2.81495 |
| 73 | L. Lenticular nucleus, putamen | 41 | L. Amygdala | 0.00537 | -2.7898 |
| 82 | R. Superior temporal gyrus | 42 | R. Amygdala | 0.005646 | -2.77337 |
| 74 | R. Lenticular nucleus, putamen | 30 | R. Insula | 0.005975 | -2.75475 |
| 57 | L. Postcentral gyrus | 41 | L. Amygdala | 0.006141 | -2.74572 |
| 41 | L. Amygdala | 33 | L. Median cingulate and paracingulate gyri | 0.00636 | -2.73413 |
| 84 | R. Temporal pole: superior temporal gyrus | 74 | R. Lenticular nucleus, putamen | 0.006381 | -2.73305 |
| 80 | R. Heschl gyrus | 73 | L. Lenticular nucleus, putamen | 0.006477 | -2.72806 |
| 83 | L. Temporal pole: superior temporal gyrus | 42 | R. Amygdala | 0.006519 | -2.72592 |
| 44 | R. Calcarine fissure and surrounding cortex | 41 | L. Amygdala | 0.006537 | -2.72502 |
| 76 | R. Lenticular nucleus, pallidum | 58 | R. Postcentral gyrus | 0.006625 | -2.72057 |

**Table S6.** Top 50 Significant FC correlations associated with frequency of SDV are presented in the table below, summarized from Figure 3. A total of 256 models were statistically significant (q < 0.0050) after FDR correction. A positive t-value indicates increased FC associated with SDV frequency, while a negative t-value indicates decreased FC associated with SDV frequency. Abbreviations: FC, functional connectivity; SDV, self-directed violence; FDR, false discovery rate

| **Label_1** | **Region_1** | **Label_2** | **Region_2** | **p_value** | **t_value** |
| --- | --- | --- | --- | --- | --- |
| 46 | R. Cuneus | 10 | R. Middle frontal gyrus, orbital part | 8.07E-05 | 3.957758 |
| 54 | R. Inferior occipital gyrus | 26 | R. Superior frontal gyrus, medial orbital | 0.00014 | 3.821511 |
| 54 | R. Inferior occipital gyrus | 9 | L. Middle frontal gyrus, orbital part | 0.000469 | 3.509202 |
| 54 | R. Inferior occipital gyrus | 6 | R. Superior frontal gyrus, orbital part | 0.000573 | 3.454517 |
| 45 | L. Cuneus | 10 | R. Middle frontal gyrus, orbital part | 0.000601 | 3.441642 |
| 46 | R. Cuneus | 7 | L. Middle frontal gyrus | 0.000693 | 3.40247 |
| 46 | R. Cuneus | 6 | R. Superior frontal gyrus, orbital part | 0.000734 | 3.386684 |
| 46 | R. Cuneus | 8 | R. Middle frontal gyrus | 0.000825 | 3.354057 |
| 45 | L. Cuneus | 9 | L. Middle frontal gyrus, orbital part | 0.001143 | 3.261643 |
| 46 | R. Cuneus | 26 | R. Superior frontal gyrus, medial orbital | 0.001172 | 3.254591 |
| 46 | R. Cuneus | 9 | L. Middle frontal gyrus, orbital part | 0.001277 | 3.229915 |
| 48 | R. Lingual gyrus | 16 | R. Inferior frontal gyrus, orbital part | 0.001307 | 3.223162 |
| 53 | L. Inferior occipital gyrus | 26 | R. Superior frontal gyrus, medial orbital | 0.001732 | 3.140873 |
| 66 | R. Angular gyrus | 46 | R. Cuneus | 0.002257 | 3.061619 |
| 47 | L. Lingual gyrus | 16 | R. Inferior frontal gyrus, orbital part | 0.002356 | 3.048705 |
| 49 | L. Superior occipital gyrus | 26 | R. Superior frontal gyrus, medial orbital | 0.002377 | 3.04604 |
| 53 | L. Inferior occipital gyrus | 9 | L. Middle frontal gyrus, orbital part | 0.00268 | 3.009471 |
| 45 | L. Cuneus | 26 | R. Superior frontal gyrus, medial orbital | 0.002767 | 2.999627 |
| 51 | L. Middle occipital gyrus | 9 | L. Middle frontal gyrus, orbital part | 0.003027 | 2.971959 |
| 44 | R. Calcarine fissure and surrounding cortex | 16 | R. Inferior frontal gyrus, orbital part | 0.003386 | 2.937035 |
| 50 | R. Superior occipital gyrus | 26 | R. Superior frontal gyrus, medial orbital | 0.003896 | 2.892866 |
| 46 | R. Cuneus | 4 | R. Superior frontal gyrus, dorsolateral | 0.004035 | 2.881767 |
| 74 | R. Lenticular nucleus, putamen | 11 | L. Inferior frontal gyrus, opercular part | 0.004211 | 2.868156 |
| 54 | R. Inferior occipital gyrus | 10 | R. Middle frontal gyrus, orbital part | 0.004455 | 2.850202 |
| 56 | R. Fusiform gyrus | 26 | R. Superior frontal gyrus, medial orbital | 0.004561 | 2.842605 |
| 49 | L. Superior occipital gyrus | 6 | R. Superior frontal gyrus, orbital part | 0.004813 | 2.825295 |
| 46 | R. Cuneus | 16 | R. Inferior frontal gyrus, orbital part | 0.004859 | 2.822216 |
| 51 | L. Middle occipital gyrus | 26 | R. Superior frontal gyrus, medial orbital | 0.004896 | 2.819788 |
| 44 | R. Calcarine fissure and surrounding cortex | 10 | R. Middle frontal gyrus, orbital part | 0.005408 | 2.787473 |
| 52 | R. Middle occipital gyrus | 26 | R. Superior frontal gyrus, medial orbital | 0.005411 | 2.787282 |
| 45 | L. Cuneus | 8 | R. Middle frontal gyrus | 0.005486 | 2.78281 |
| 45 | L. Cuneus | 6 | R. Superior frontal gyrus, orbital part | 0.005523 | 2.780598 |
| 48 | R. Lingual gyrus | 10 | R. Middle frontal gyrus, orbital part | 0.005948 | 2.756253 |
| 43 | L. Calcarine fissure and surrounding cortex | 16 | R. Inferior frontal gyrus, orbital part | 0.005973 | 2.754861 |
| 48 | R. Lingual gyrus | 9 | L. Middle frontal gyrus, orbital part | 0.006341 | 2.735115 |
| 54 | R. Inferior occipital gyrus | 5 | L. Superior frontal gyrus, orbital part | 0.006504 | 2.726684 |
| 49 | L. Superior occipital gyrus | 10 | R. Middle frontal gyrus, orbital part | 0.006784 | 2.712633 |
| 43 | L. Calcarine fissure and surrounding cortex | 10 | R. Middle frontal gyrus, orbital part | 0.006944 | 2.704882 |
| 49 | L. Superior occipital gyrus | 9 | L. Middle frontal gyrus, orbital part | 0.007054 | 2.699587 |
| 45 | L. Cuneus | 16 | R. Inferior frontal gyrus, orbital part | 0.007149 | 2.69511 |
| 49 | L. Superior occipital gyrus | 8 | R. Middle frontal gyrus | 0.007433 | 2.682034 |
| 51 | L. Middle occipital gyrus | 31 | L. Anterior cingulate and paracingulate gyri | 0.007511 | 2.678514 |
| 87 | L. Temporal pole: middle temporal gyrus | 15 | L. Inferior frontal gyrus, orbital part | 0.007579 | 2.675462 |
| 54 | R. Inferior occipital gyrus | 25 | L. Superior frontal gyrus, medial orbital | 0.007698 | 2.670204 |
| 26 | R. Superior frontal gyrus, medial orbital | 3 | L. Superior frontal gyrus, dorsolateral | 0.007905 | 2.661198 |
| 53 | L. Inferior occipital gyrus | 7 | L. Middle frontal gyrus | 0.008178 | 2.649673 |
| 53 | L. Inferior occipital gyrus | 6 | R. Superior frontal gyrus, orbital part | 0.008467 | 2.637855 |
| 51 | L. Middle occipital gyrus | 6 | R. Superior frontal gyrus, orbital part | 0.008591 | 2.632885 |
| 70 | R. Paracentral lobule | 65 | L. Angular gyrus | 0.008959 | 2.618486 |
| 65 | L. Angular gyrus | 46 | R. Cuneus | 0.009039 | 2.615453 |

**Table S7.** Top 50 significant FC correlations associated with interval of SDV are presented in the table below, summarized from Figure 4. A total of 155 models were statistically significant after FDR correction (q < 0.0498). Positive t-values indicate increased FC correlation associated with SDV interval, while a negative t-value indicates decreased FC correlation associated with SDV interval. Abbreviations: FC, functional connectivity; SDV, self-directed violence; FDR, false discovery rate

| **Label_1** | **Region_1** | **Label_2** | **Region_2** | **p_value** | **t_value** |
| --- | --- | --- | --- | --- | --- |
| 48 | R. Lingual gyrus | 43 | L. Calcarine fissure and surrounding cortex | 0.000823 | -3.35463 |
| 90 | R. Inferior temporal gyrus | 32 | R. Anterior cingulate and paracingulate gyri | 0.000878 | -3.33647 |
| 27 | L. Gyrus rectus | 24 | R. Superior frontal gyrus, medial | 0.001021 | -3.29405 |
| 64 | R. Supramarginal gyrus | 34 | R. Median cingulate and paracingulate gyri | 0.001365 | -3.21062 |
| 68 | R. Precuneus | 60 | R. Superior parietal gyrus | 0.001896 | -3.11398 |
| 90 | R. Inferior temporal gyrus | 4 | R. Superior frontal gyrus, dorsolateral | 0.002783 | -2.99787 |
| 86 | R. Middle temporal gyrus | 8 | R. Middle frontal gyrus | 0.002962 | -2.97863 |
| 64 | R. Supramarginal gyrus | 21 | L. Olfactory cortex | 0.003406 | -2.93519 |
| 6 | R. Superior frontal gyrus, orbital part | 2 | R. Precentral gyrus | 0.003691 | -2.91001 |
| 87 | L. Temporal pole: middle temporal gyrus | 8 | R. Middle frontal gyrus | 0.004289 | -2.86231 |
| 90 | R. Inferior temporal gyrus | 24 | R. Superior frontal gyrus, medial | 0.005127 | -2.80483 |
| 89 | L. Inferior temporal gyrus | 65 | L. Angular gyrus | 0.005172 | -2.802 |
| 68 | R. Precuneus | 12 | R. Inferior frontal gyrus, opercular part | 0.006383 | -2.73293 |
| 86 | R. Middle temporal gyrus | 10 | R. Middle frontal gyrus, orbital part | 0.006479 | -2.72796 |
| 27 | L. Gyrus rectus | 11 | L. Inferior frontal gyrus, opercular part | 0.006884 | -2.70778 |
| 89 | L. Inferior temporal gyrus | 3 | L. Superior frontal gyrus, dorsolateral | 0.007269 | -2.68953 |
| 86 | R. Middle temporal gyrus | 6 | R. Superior frontal gyrus, orbital part | 0.007361 | -2.68531 |
| 32 | R. Anterior cingulate and paracingulate gyri | 27 | L. Gyrus rectus | 0.008134 | -2.65151 |
| 68 | R. Precuneus | 2 | R. Precentral gyrus | 0.008274 | -2.64569 |
| 27 | L. Gyrus rectus | 16 | R. Inferior frontal gyrus, orbital part | 0.008711 | -2.62814 |
| 35 | L. Posterior cingulate gyrus | 11 | L. Inferior frontal gyrus, opercular part | 0.008916 | -2.62016 |
| 88 | R. Temporal pole: middle temporal gyrus | 10 | R. Middle frontal gyrus, orbital part | 0.009675 | -2.59199 |
| 25 | L. Superior frontal gyrus, medial orbital | 11 | L. Inferior frontal gyrus, opercular part | 0.010522 | -2.5628 |
| 90 | R. Inferior temporal gyrus | 35 | L. Posterior cingulate gyrus | 0.010704 | -2.55679 |
| 84 | R. Temporal pole: superior temporal gyrus | 34 | R. Median cingulate and paracingulate gyri | 0.011442 | -2.53336 |
| 87 | L. Temporal pole: middle temporal gyrus | 7 | L. Middle frontal gyrus | 0.011628 | -2.52768 |
| 27 | L. Gyrus rectus | 10 | R. Middle frontal gyrus, orbital part | 0.011806 | -2.52229 |
| 89 | L. Inferior temporal gyrus | 7 | L. Middle frontal gyrus | 0.01227 | -2.50862 |
| 25 | L. Superior frontal gyrus, medial orbital | 24 | R. Superior frontal gyrus, medial | 0.012738 | -2.49529 |
| 90 | R. Inferior temporal gyrus | 26 | R. Superior frontal gyrus, medial orbital | 0.012965 | -2.48899 |
| 87 | L. Temporal pole: middle temporal gyrus | 66 | R. Angular gyrus | 0.013177 | -2.48316 |
| 82 | R. Superior temporal gyrus | 65 | L. Angular gyrus | 0.013366 | 2.478083 |
| 50 | R. Superior occipital gyrus | 23 | L. Superior frontal gyrus, medial | 0.013366 | 2.478075 |
| 66 | R. Angular gyrus | 3 | L. Superior frontal gyrus, dorsolateral | 0.013722 | -2.46862 |
| 87 | L. Temporal pole: middle temporal gyrus | 10 | R. Middle frontal gyrus, orbital part | 0.013912 | -2.46367 |
| 8 | R. Middle frontal gyrus | 2 | R. Precentral gyrus | 0.014407 | -2.45103 |
| 66 | R. Angular gyrus | 46 | R. Cuneus | 0.015554 | 2.423175 |
| 11 | L. Inferior frontal gyrus, opercular part | 5 | L. Superior frontal gyrus, orbital part | 0.015842 | -2.41646 |
| 66 | R. Angular gyrus | 27 | L. Gyrus rectus | 0.016117 | -2.41015 |
| 64 | R. Supramarginal gyrus | 22 | R. Olfactory cortex | 0.016125 | -2.40997 |
| 27 | L. Gyrus rectus | 8 | R. Middle frontal gyrus | 0.016512 | -2.40125 |
| 71 | L. Caudate nucleus | 24 | R. Superior frontal gyrus, medial | 0.016739 | -2.39623 |
| 90 | R. Inferior temporal gyrus | 31 | L. Anterior cingulate and paracingulate gyri | 0.017322 | -2.38358 |
| 87 | L. Temporal pole: middle temporal gyrus | 71 | L. Caudate nucleus | 0.017709 | -2.37539 |
| 27 | L. Gyrus rectus | 15 | L. Inferior frontal gyrus, orbital part | 0.017995 | -2.36945 |
| 65 | L. Angular gyrus | 46 | R. Cuneus | 0.018287 | 2.363446 |
| 68 | R. Precuneus | 63 | L. Supramarginal gyrus | 0.018395 | -2.36126 |
| 71 | L. Caudate nucleus | 35 | L. Posterior cingulate gyrus | 0.019391 | -2.34153 |
| 35 | L. Posterior cingulate gyrus | 10 | R. Middle frontal gyrus, orbital part | 0.019527 | -2.33892 |
| 77 | L. Thalamus | 24 | R. Superior frontal gyrus, medial | 0.020124 | -2.32757 |

Alfaro-Almagro, F., Jenkinson, M., Bangerter, N. K., Andersson, J. L. R., Griffanti, L., Douaud, G., Sotiropoulos, S. N., Jbabdi, S., Hernandez-Fernandez, M., Vallee, E., Vidaurre, D., Webster, M., McCarthy, P., Rorden, C., Daducci, A., Alexander, D. C., Zhang, H., Dragonu, I., Matthews, P. M., . . . Smith, S. M. (2018). Image processing and Quality Control for the first 10,000 brain imaging datasets from UK Biobank. *Neuroimage*, *166*, 400-424. <https://doi.org/10.1016/j.neuroimage.2017.10.034>

Crosby, A., Ortega, L., & Melanson, C. (2011). Self-directed violence surveillance; uniform definitions and recommended data elements.

Tang, L. (2018). Next-generation peptide sequencing. *Nat Methods*, *15*(12), 997. <https://doi.org/10.1038/s41592-018-0240-7>
